# Characteristics and transmission dynamics of COVID-19 in healthcare workers at a London teaching hospital

**DOI:** 10.1101/2020.07.10.20149237

**Authors:** Charlotte Zheng, Nema Hafezi-Bakhtiari, Victoria Cooper, Harriet Davidson, Maximillian Habibi, Peter Riley, Aodhan Breathnach

## Abstract

**Background:** Healthcare worker (HCW) associated COVID-19 is of global concern due to the potential for nosocomial spread and depletion of staff numbers. However, the literature on transmission routes and risk factors for COVID-19 in HCWs is limited.

**Aim:** To examine the characteristics and transmission dynamics of SARS-CoV-2 in HCWs in a university teaching hospital in London, UK.

**Methods:** Staff records and virology testing results were combined to identify staff sickness and COVID-19 rates from March to April 2020. Comparisons were made with staff professional groups, department of work and ethnicity. Analysis was performed using Microsoft Excel™.

**Findings:** COVID-19 rates in our HCWs largely rose and declined in parallel with the number of community cases. White and non-white ethnic groups among our HCWs had similar rates of infection. Clinical staff had a higher rate of laboratory-confirmed COVID-19 than non-clinical staff, but total sickness rates were similar. Doctors had the highest rate of infection, but took the fewest sickness days. Critical Care had lower rates than the Emergency Department (ED), but rates in the ED declined once all staff were advised to use Personal Protective Equipment (PPE).

**Conclusion:** These findings show that sustained transmission of SARS-CoV-2 among our hospital staff did not occur, beyond the community outbreak, even in the absence of strict infection control measures in non-clinical areas. The results also suggest that current PPE is effective when used appropriately. In addition, our findings emphasise the importance of testing both clinical and non-clinical staff groups during a pandemic.

## Introduction

Coronavirus disease 2019 (COVID-19) in healthcare workers (HCWs) has caused understandable concern because of the risk of infection from patients, the impact on staffing levels and the potential for hospital staff to become vectors for onward transmission. Reports of worse outcomes in BAME (Black, Asian and other Minority Ethnic) groups and debates about personal protective equipment (PPE) have heightened these concerns (1, 2). China, Italy and the USA have reported HCW infection rates of up to 3.8%, 10% and 19% respectively with fatality rates of up to 1.2% (3-5). Current literature for the UK is limited and is restricted to data from short time-frames with little detail on transmission dynamics, and inter-departmental and inter-specialty differences (6, 7).

On 18^th^ March 2020 we started testing staff for acute infection in our own institution (a London teaching hospital, employing 8738 staff). We were aware of significant numbers of infections, and sadly four staff members from clinical and non-clinical settings have died. By analysing our staff testing data, we hoped to identify patterns of transmission and risk factors for disease acquisition.

## Methods

Staff testing started a week after the ‘surge’ in COVID-19 admissions began. Criteria for testing changed over time due to guidance from NHS England, increases in testing capacity and a growing awareness of the range of staff being infected. Initial testing focused on front-line clinical staff, especially from the Emergency Department (ED) and Intensive Care Unit (ICU). This was gradually expanded to other clinical staff, then to all staff, and finally to contractors (including cleaning and catering staff). Symptomatic staff were referred by their line managers to a drive-through testing pod. A combined nose and throat swab was taken for SARS-CoV-2 real-time PCR. We initially used the E and S gene target assay (Realstar®, Altona Diagnostics]), and later replaced this with the ORF1a/b and E gene target assay (Cobas® SARS-CoV-2 assay, Roche). Occupational health data and staff records were combined to identify proven COVID-19 and sickness rates from March to April 2020 and analysed using Microsoft Excel™. Staff were categorised as clinical (nurses, doctors, allied health professionals, healthcare assistants) or non-clinical (administrative, non-patient facing specialties e.g. laboratory scientists, housekeeping, estates and facilities etc.).

## Results

1045 hospital staff were tested for SARS-CoV-2 infection by PCR, of which staff roles could be identified in 958 (92%), comprising 11% of overall staff numbers. SARS-CoV-2 was detected in 498 (52%). The proportion of male staff in the hospital both attending for testing and testing positive was higher than in females: 13% versus 10% (p=0.002) and 7% versus 5% (p=0.0006) respectively. Ethnicity data was available for 778 staff. The proportions of white and BAME staff in the hospital attending for COVID-19 testing and subsequently testing positive were broadly similar (*Figure 1*). However, there were differences within the BAME groups; in particular, a lower proportion of Black/Black British staff attended for testing and tested positive. This may be related to the differences noted in the representation of different ethnic groups in different professional groups. In particular just 2% of the medical/dental workforce is comprised of Black/Black British staff but they make up 9% and 16% of the nursing and HCA staff groups respectively.

**Figure 1.**
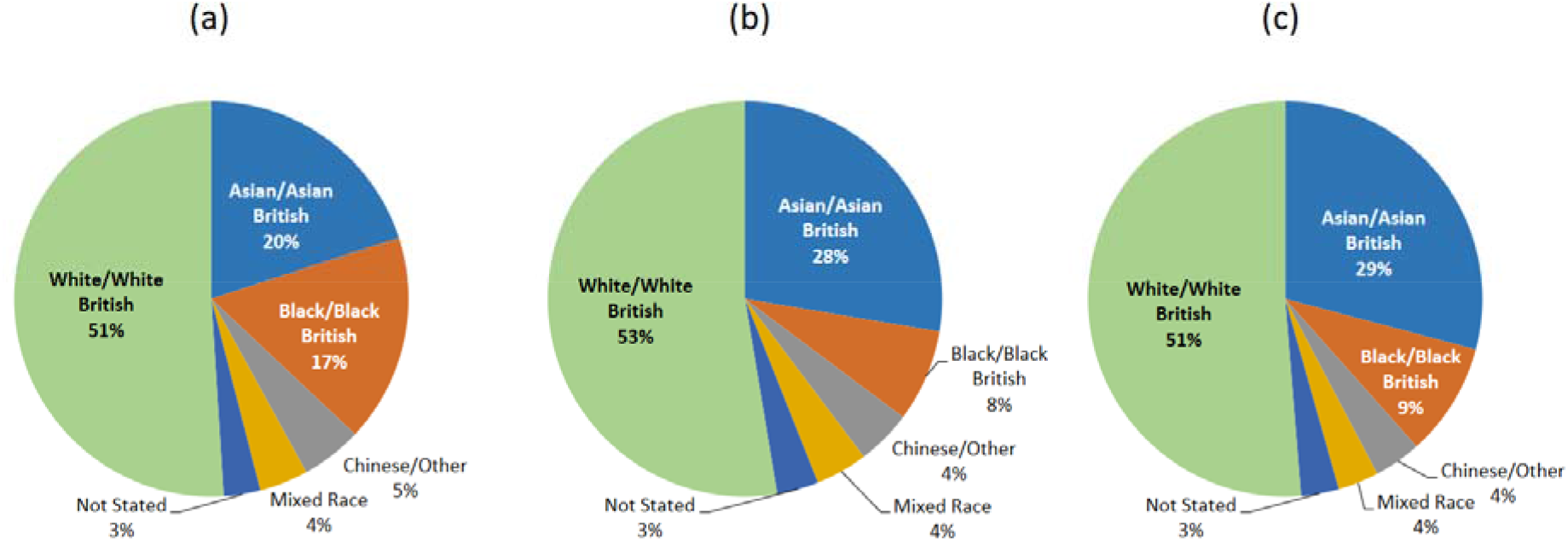
Proportion of ethnic groups in (a) All HCWs in this hospital (b) HCWs who attended for testing (c) HCWs who tested positive for SARS-CoV-2

Infections occurred in all staff groups and in all departments in the hospital. The epidemic curves for new admissions of COVID-19 patients, COVID-19 positive staff and staff sickness episodes are shown in *Figure 2*. The curves coincide closely, although absence due to illness (from any cause) in clinical staff peaked a week after that in non-clinical staff, and coincided with the peak of COVID-19 patient admissions. A possible second smaller peak in staff sickness is observed about a week after testing for all clinical and non-clinical staff was introduced. The peak of confirmed staff COVID-19 actually occurred a week before that of patient admissions.

**Figure 2.**
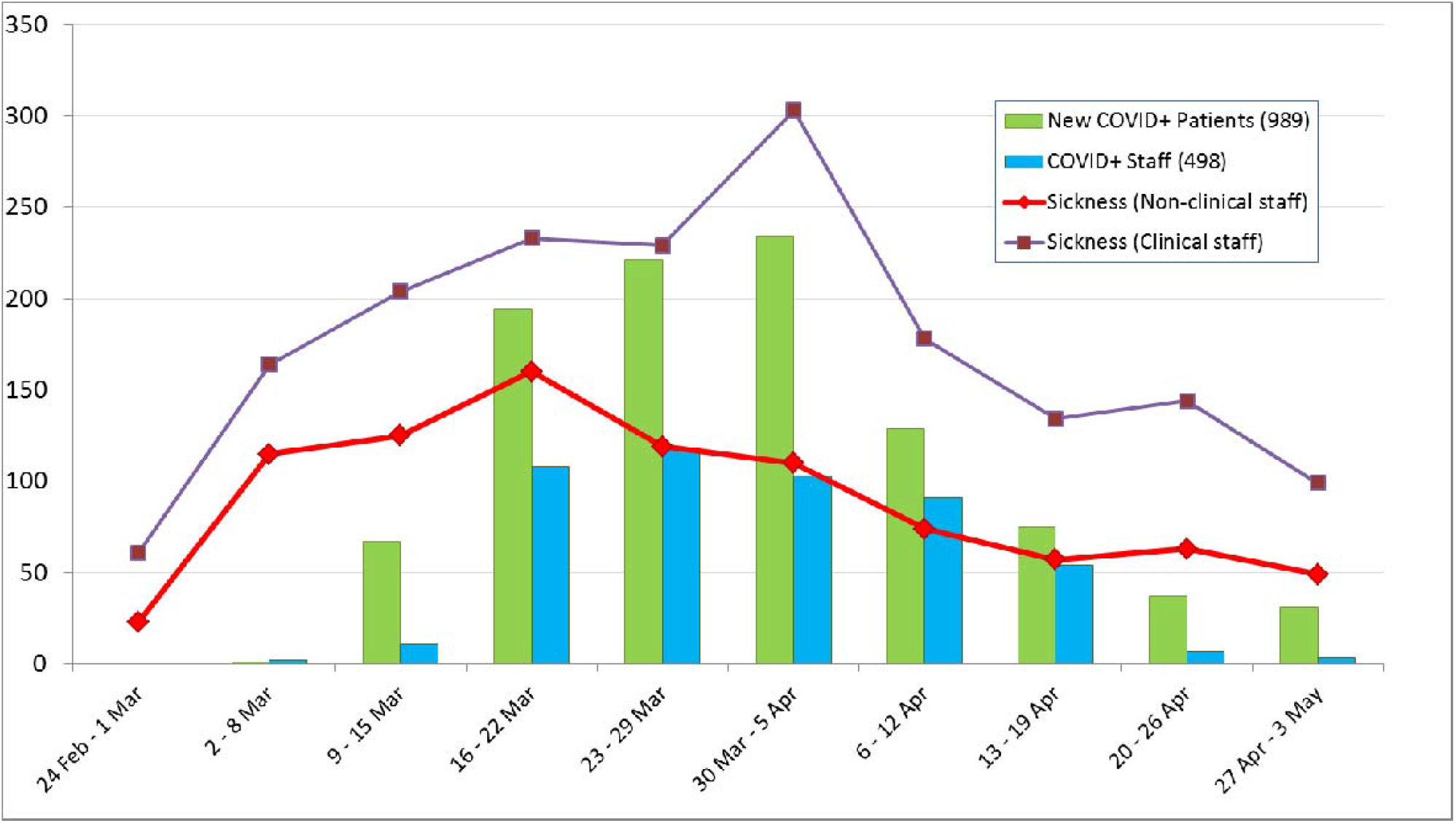
Epidemic curve showing weekly numbers of patients admitted, COVID-19 in staff, and total sickness episodes for clinical and non-clinical staff between 24th February and 3rd May 2020. The dates for COVID-19 positive staff refer to the onset of illness, not the testing date. Testing of inpatients without a travel history began on 12^th^ March. The official staff testing programme began on 18 March. The UK national lockdown began on 23^rd^ March.

A higher proportion of clinical staff tested positive for SARS-CoV-2 over the study period, compared to non-clinical staff groups (7% and 3% respectively). Doctors had the highest rate of proven COVID-19 at 11%, followed by nurses at 7% and healthcare assistants (HCAs) at 6%, as shown in *Figure 3*. Because clinical staff were initially prioritised for testing, we also examined staff sickness records. Total staff illness episodes were proportionately similar in both clinical and non-clinical groups, despite clinical staff being more likely to attribute their illness to COVID-19 (*Table I*). 30% of staff had an episode of sick leave in March and April, accounting for 29,862 days lost, a 74% increase in sickness levels compared with the same time period in the previous year. Despite having the highest rate of proven COVID-19; doctors had the lowest rate of overall sickness absence compared to other staff groups, at a mean of 1.4 days.

**Figure 3.**
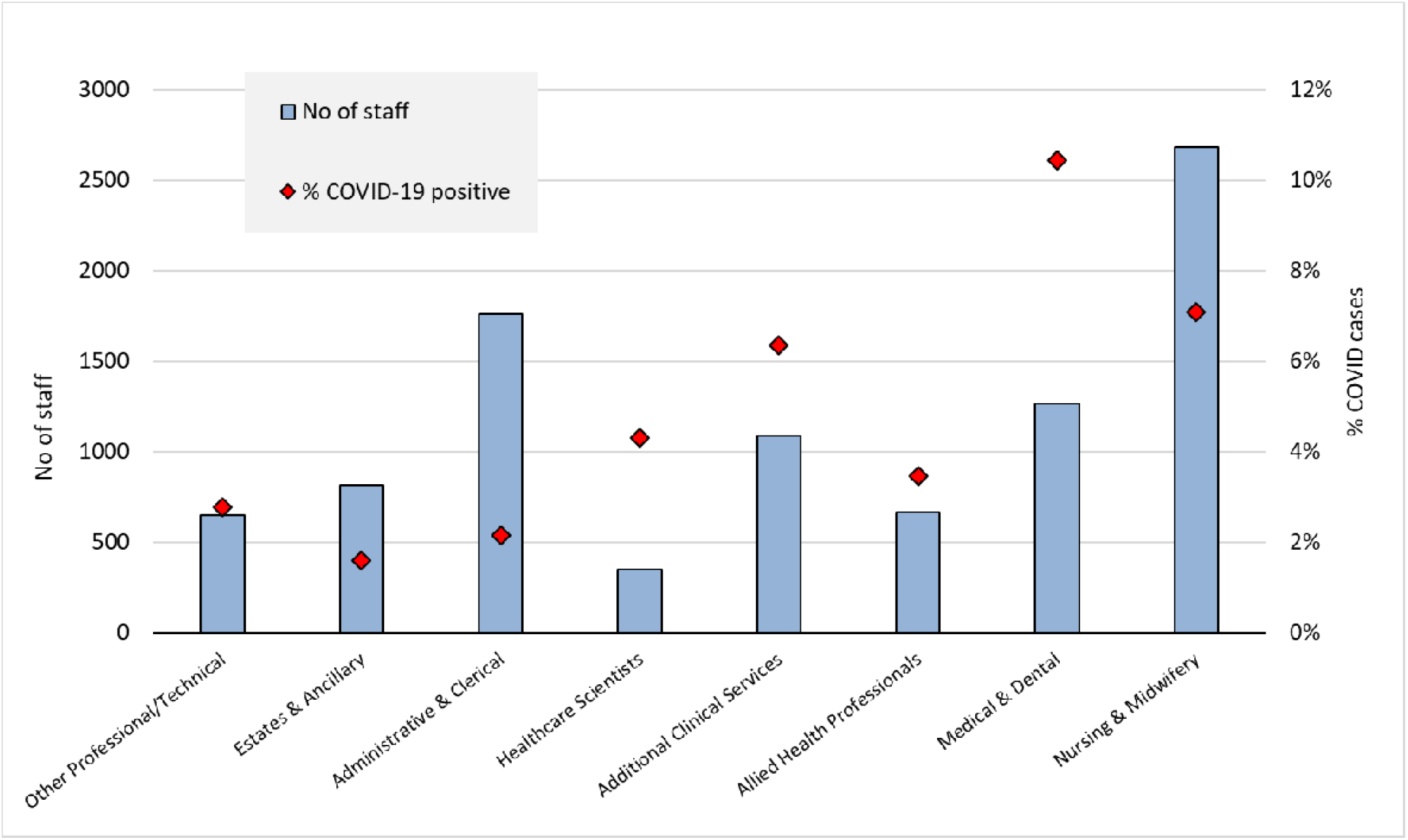
Numbers of staff in different professional groups, and proportion of those groups diagnosed with COVID-19. The ‘Additional Clinical Services’ group consists mostly of healthcare assistants.

**Table I.**
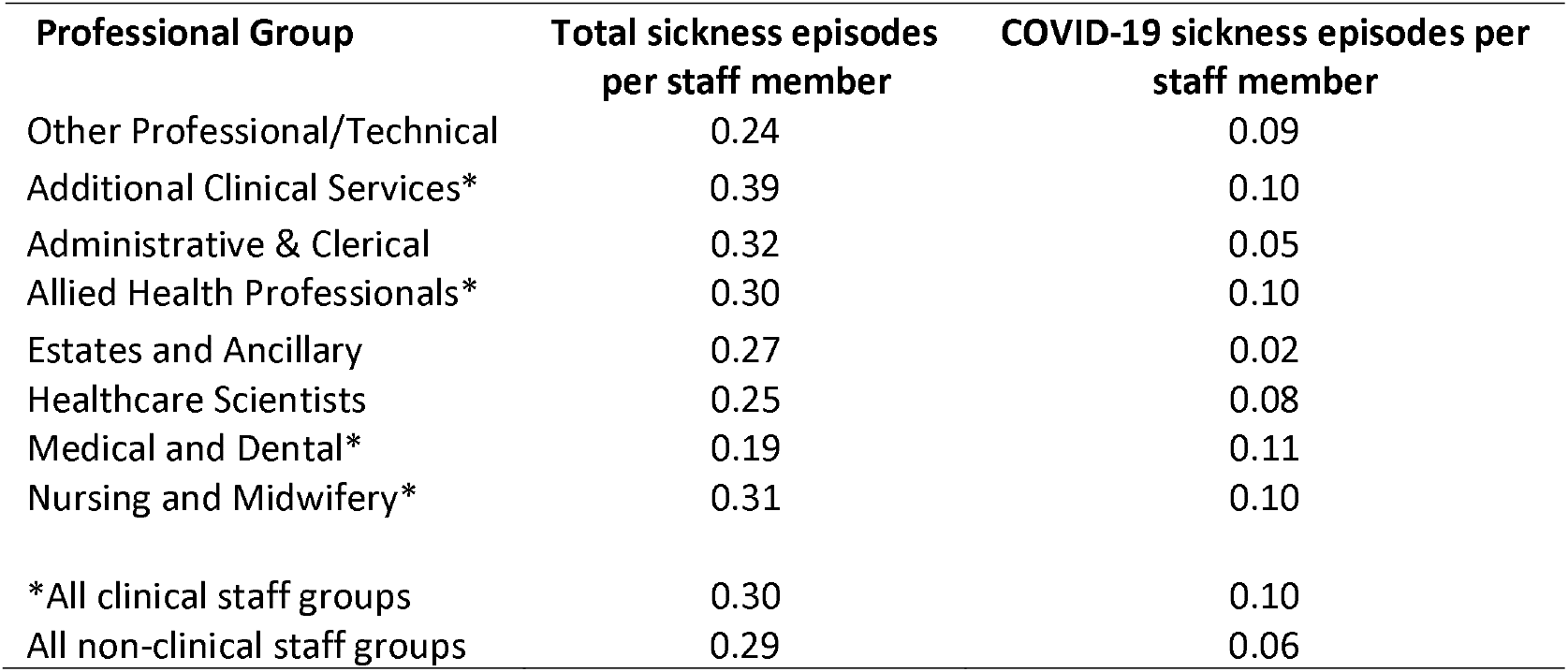
Episodes of COVID-19 specific and overall sickness in different staff groups, during March and April 2020

Rates of infection varied widely between clinical departments (examples of rates in selected departments given in *Table II*). This data records the departments that staff are formally assigned to, but many general medical and surgical staff were rapidly assigned to newly-designated COVID-19 wards and intensive care units (ICUs) during these months. Anaesthetists and theatre staff tended to be assigned specifically to COVID-19 ICUs. Front-line services – emergency and acute medicine – had considerably higher rates of proven COVID-19 than ICU and theatre staff. Confirmed COVID-19 infections also peaked earlier in the ED and acute medicine compared to other specialties (*Figure 3*). Of note, staff infections in the ED decreased after the introduction of universal PPE in the department from 28^th^ March. In contrast, confirmed infections peaked later in locum/bank staff.

**Table II.** Rates of COVID-19 in selected hospital directorates

| Directorate | Number of staff | Number diagnosed with COVID-19 | % COVID-19 positive |
| --- | --- | --- | --- |
| Emergency Medicine | 231 | 40 | 17.3% |
| Acute Medicine | 538 | 56 | 10.4% |
| Cardiothoracic | 195 | 18 | 9.2% |
| Cardiology | 271 | 24 | 8.9% |
| Renal, Oncology & Palliative Medicine | 355 | 31 | 8.7% |
| Specialist Medicine (including Infectious Disease & Respiratory) | 385 | 33 | 8.6% |
| Surgery | 718 | 38 | 5.3% |
| Neurosciences | 616 | 29 | 4.7% |
| Pathology | 493 | 20 | 4.1% |
| Diagnostics (inc Radiology) | 425 | 17 | 4.0% |
| Estates & Facilities | 340 | 13 | 3.8% |
| Anaesthetics and Theatres | 645 | 23 | 3.6% |
| Therapies | 309 | 11 | 3.6% |
| Outpatients | 440 | 12 | 2.7% |
| Children's Services | 742 | 20 | 2.7% |
| Critical Care | 428 | 11 | 2.6% |
| Women's Services. | 414 | 10 | 2.4% |
| Pharmacy Dir. | 268 | 5 | 1.9% |

## Discussion

Limitations of our data include a lack of information on disease severity and clinical outcomes as well as the effect of staff redeployment to COVID-19 wards and ICUs. We also have less data available for contracted services, which includes many domestic and cleaning staff. The true rate of COVID-19 in different staff groups may be masked by selective and changing testing criteria. This was addressed by analysing overall staff sickness episodes.

When the COVID-19 pandemic began, there was global concern about the risks to HCWs and the adequacy of PPE. Front-line clinical staff were perceived to be at greatest risk, and this (along with concerns about diagnostic capacity) informed the initial staff testing strategy. However, the matching epidemic curves of proven staff and patient infections along with the large numbers of infections in non-clinical staff supports a community source for a significant proportion of staff. Nevertheless, the delayed peak in clinical staff sickness episodes cannot be ignored. The most plausible explanation is that at least some of the staff infections are related to patient exposure, with some transmission within individual clinical departments. Department-specific data does support a hypothesis of some localised clusters of infection (*Table II*). This is not surprising given viral infectivity and necessary close contact of staff in a busy work environment. The possible second smaller peak in staff sickness may represent increased detection due to widening of criteria of testing to all staff groups. Nonetheless, the fact that staff COVID-19 and sickness rates decreased rapidly through April, in line with the decrease in COVID-19 patient admissions, suggests that sustained hospital transmission did not occur, despite the localised clusters, and despite the ongoing proximity of staff to each other and to inpatients with COVID-19. This is perhaps relevant to the current debate about what mandatory measures for staff are necessary to prevent and manage possible future epidemics of COVID-19 in hospitals.

We found no evidence of increased acquisition of COVID-19 among BAME staff, as the rates reflected overall staff proportions. As noted above, however, we were unable to gather data on disease severity or on the proportion of staff admitted to hospital. The under-representation of Black/Black British staff attending for testing was surprising, and may have been due to the different representation of ethnic minorities in particular staff groups leading to differential access to testing, especially early in the local epidemic.

The testing data and overall sickness rates gave conflicting results for clinical and non-clinical staff groups. This may partly be because non-clinical staff had reduced access to testing. It may also reflect varying pressures around taking sick leave – particularly among doctors, who had the least documented sick leave, despite having the most proven COVID-19. Infected but mildly ill staff members may be tempted to continue working (especially if their roles are highly specialised and cannot be easily covered by a colleague) thus posing an on-going transmission risk (8). It is important to provide access to testing across all hospital staff groups as despite being denoted “non-clinical”, many employees (eg domestic staff) work in clinical areas and access communal areas. Testing all staff groups has crucial infection control implications as it allows detection of infectious workers, and can enable non-infectious colleagues to return to work. Furthermore, unequal access to testing may lead excluded staff groups to feel undervalued.

Earlier peaks in staff COVID-19 infections in acute medicine and the ED compared to other specialties are likely to reflect those services being the first point of contact for patients but also that testing was prioritised for frontline services early on in the epidemic. The later peak in locum/bank staff may be partially explained by these staff being employed later on in the pandemic as demand for staffing increased due to the surge in COVID-19 admissions. They may have also had increased exposure from working in different healthcare settings and institutions.

Amongst HCWs who were consistently able to access testing, rates of positive tests are higher for ED and acute medicine than for ICU. ICU is often regarded as the highest risk working environment, with a higher frequency of aerosol generating procedures. This finding is consistent with national statistics around deaths in HCWs (9). It is tempting to attribute this simply to the enhanced PPE that is routine in ICU, but other possibilities must also be considered. ED has a more hectic and cramped working environment; many COVID-19 cases (especially early in the surge) would be initially unrecognised; staff would alternate working in COVID-19 and non-COVID-19 designated areas. Patients attending ED may also be earlier in their illness, with higher levels of virus shedding (10), and generally are not ventilated. When PPE was mandated for staff in all clinical areas of ED, the rates of COVID-19 dropped (*Figure 4*).

**Figure 4.**
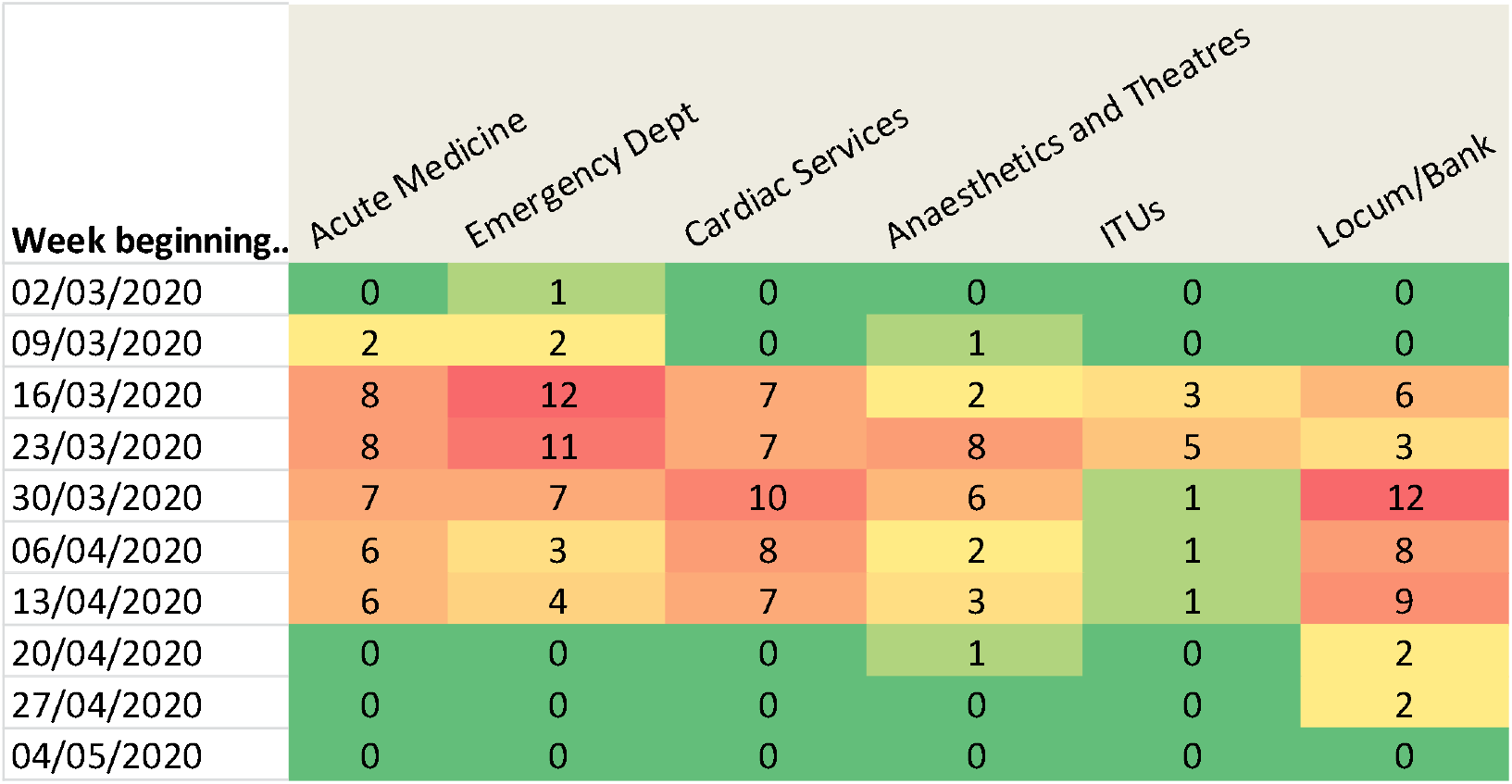
Heat map showing weekly numbers of confirmed COVID-19 in selected clinical departments from March to April 2020.

## Conclusion

These results have shown that all staff groups are at risk of COVID-19 with rates generally reflecting community patterns of transmission, although front-line clinical staff may be at increased risk. Sustained spread of COVID-19 among staff, beyond the peak in community cases, did not occur. Differences in rates of confirmed infections amongst hospital departments and professional groups may in part be due to differential access to testing: thus it is important that in future there is equality of access to testing for all staff, including those whose employment may be contracted out to the private sector such as domestic staff. There needs to be early recognition of possible cases in acute settings, with an emphasis on universal application of diligent basic hygiene and PPE. There also needs to be clarity about when staff should go off sick, with sufficient support to ensure work is cross-covered safely, so that staff do not continue to work with mild symptoms and risk transmitting COVID-19 to other colleagues or patients.

## Data Availability

Data is available on request from the corresponding author.

